# Longitudinal tau PET using [^18^F]flortaucipir: Comparison of (semi)quantitative parameters

**DOI:** 10.1101/2021.08.27.21262740

**Authors:** Denise Visser, Hayel Tuncel, Rik Ossenkoppele, Maqsood Yaqub, Emma E Wolters, Tessa Timmers, Emma Weltings, Emma M Coomans, Marijke den Hollander, Wiesje M van der Flier, Bart NM van Berckel, Sandeep SV Golla

**Author notes:** Corresponding author: Denise Visser, Department of Radiology & Nuclear Medicine, Amsterdam University Medical Centers, location VUmc, Amsterdam, The Netherlands, P.O. Box 7057, 1007 MB Amsterdam, The Netherlands. both authors contributed equally (Denise Visser, Hayel Tuncel). Financial disclosure: This research was funded by a ZonMW Memorabel grant.

## Abstract

**Purpose:** Semi-quantitative PET measures can be affected by changes in blood flow, whereas quantitative measures are not. The aim of the study was to compare semi-quantitative(SUVr) and quantitative(R_1_, BP_ND_) parameters of longitudinal tau PET scans with [^18^F]flortaucipir, with respect to changes in blood flow.

**Methods:** Subjects with subjective cognitive decline(SCD; n=38) and Alzheimer’s disease(AD) patients(n=24) underwent baseline(BL) and 2-year follow-up(FU) dynamic [^18^F]flortaucipir PET scans. BP_ND_ and R_1_ were estimated using RPM and SUVr_(80-100min)_ was calculated (cerebellar gray as reference). For each region-of-interest ((trans)entorhinal, limbic and neocortical) and parameter, %change was calculated. Regional SUVrs were compared to corresponding DVR (= BP_ND_+1) using paired T-tests. Additionally, simulations were performed to model effects of flow changes on BP_ND_ and SUVr in different binding categories. Thereafter, %bias for SUVr with respect to underlying binding and flow were evaluated.

**Results:** In SCD, there was a difference between %change in the (trans)entorhinal ROI (DVR 2.56% vs SUVr 1.85%) only. In AD, a difference was found in the limbic ROI (DVR 6.61% vs SUVr 7.52%) only. R_1_ changes were small(+0.7% in SCD and -1.6% in AD). Simulations illustrated with increasing flow a decreased %bias for SUVr in low binding conditions, whereas a slightly increased bias was observed in high binding conditions.

**Conclusion:** SUVr provided an accurate estimate of specific binding for [^18^F]flortaucipir over a two-year follow-up. However, simulations showed that flow changes can affect [^18^F]flortaucipir SUVr, hence DVR/BP_ND_ should be preferred in more advanced disease stages and/or conditions that could induce significant flow changes like pharmacotherapeutic interventions.

## INTRODUCTION

To accurately assess longitudinal changes or the effect of potential therapeutic interventions on tau accumulation (either as a direct target or downstream measure) *in vivo* tau PET can be used. Several tau PET tracers became available over the years, of which [^18^F]flortaucipir (previously known as [^18^F]T-807 or [^18^F]AV-1451) is most frequently used, and the only tau PET tracer (recently) approved by the FDA[1-5]. PET images can be acquired using static or dynamic scanning protocols. Static scanning protocols consist of tracer injection, followed by a short PET acquisition after a certain amount of time. Semi-quantitative parameters such as standardized uptake value ratio (SUVr) can be derived from a static PET scan. With dynamic scanning protocols, on the other hand, the PET acquisition starts simultaneously with tracer injection, and often lasts longer (e.g. 100 or 130 min for [^18^F]flortaucipir). Although the resulting scan duration is longer, parameters derived from a dynamic PET scan, such as distribution volume ratio (DVR) or binding potential (BP_ND_), are fully quantitative and overall more accurate[6, 7].

Semi-quantitative measures have the advantage of practical applicability and relative computational simplicity[2-5]. However, studies involving dynamic imaging not only provide more accurate parameters with respect to quantification of target of interest, but also provide estimates for relative tracer delivery (R_1_), a proxy for relative cerebral blood flow (rCBF)[7-11]. Blood flow changes can occur over time in AD due to disease progression or drug intervention. Monitoring these flow changes is important, since studies have shown that measuring longitudinal changes using semi-quantitative measures (SUVr) could be biased as they are affected by changes in blood flow, whereas quantitative measures (BP_ND_) are not affected by these blood flow changes[6, 12]. For these reasons, dynamic acquisition is preferred for longitudinal assessment. However, the above mentioned studies were performed using tracers for amyloid pathology, and for [^18^F]flortaucipir this has not been investigated yet.

Therefore, with this study we aim to compare semi-quantitative (SUVr) and quantitative (DVR/BP_ND_) parameters for [^18^F]flortaucipir PET in a longitudinal setting. First, we will do this based on a two-year follow-up observational study, in which (relative) cerebral blood flow changes (as measured with R_1_) are expected to be small. Secondly, we will use simulations to investigate how large(r) changes in (relative) cerebral blood flow (as might be induced by pharmacotherapeutic interventions) affect SUVr and DVR/BP_ND_.

## METHODS

### Participants

We included 62 subjects from the Amsterdam Dementia Cohort[13, 14], of whom 38 were cognitively normal with subjective cognitive decline (SCD) and 24 were cognitively impaired with either a diagnosis of mild cognitive impairment (MCI) due to AD[15] (n=4) or probable AD dementia[16] (n=20). The MCI due to AD and AD dementia subjects were grouped into one MCI/AD group.

All subjects underwent a standardized dementia screening, including medical history, extensive neuropsychological assessment, physical and neurological examination, lumbar puncture (LP), blood tests, electroencephalography and brain magnetic resonance imaging (MRI)[13, 14]. Diagnosis was established by consensus in a multidisciplinary meeting and met criteria according to the National Institute on Aging and Alzheimer’s Association (NIA-AA)[14, 17]. The SCD diagnosis was established based on self-reported cognitive complaints, while all clinical and cognitive investigations were normal[18]. Twelve out of 38 SCD subjects were classified as amyloid positive as evidenced by substantial Aß pathology after SUVr50-70min [^18^F]florbetapir Aß-PET scan visual assessment[19]. All MCI/AD patients were classified as amyloid positive as evidenced by CSF biomarkers for AD (i.e. CSF Aß1-42 < 813 ng/L[20]) and/or a positive Aß-PET ([^18^F]PiB or [^18^F]florbetaben) scan by visual assessment[21, 22].

Exclusion criteria for all participants were (1) non-AD dementia diagnosis, (2) significant cerebrovascular disease as assessed by MRI, (3) major traumatic brain injury, (4) major psychiatric or neurological disorders (other than AD), (5) recent substance abuse. The study protocol was approved by the Medical Ethics Review Committee of the Amsterdam UMC VU Medical center. All patients provided written informed consent prior to study participation.

### Imaging

All subjects underwent two dynamic [^18^F]flortaucipir PET scans, acquired on a Philips Ingenuity TF-64 PET/CT scanner, with a time period of 2.1±0.3 (SCD) or 2.2±0.3 (AD) years between both scanning sessions. For SCD subjects, each (130 min) scanning session consisted of two dynamic PET scans of 60 and 50 minutes respectively, with a 20 minute break in between[11, 23]. Details are described elsewhere[23]. In short, the first 60 minute dynamic scan started simultaneously with a bolus injection of 238±12 MBq [^18^F]flortaucipir (injected mass 1.1±0.9 µg). The second PET scan was co-registered to the first dynamic PET scan using Vinci (Volume Imaging in Neurological Imaging) software[24] and PET list mode data were rebinned into a total of 29 frames (1×15, 3×5, 3×10, 4×60, 2×150, 2×300, 4×600 and 10×300 seconds).

For AD subjects each (100min) scanning session consisted of two dynamic PET scans of 30 and 20 minutes respectively, with a 50 minute break in between[25]. Details are described elsewhere[25]. In short, the first 30 minute dynamic scan started simultaneously with a bolus injection 236±11 MBq [^18^F]flortaucipir (injected mass 1.1±0.8 µg). The second PET scan was co-registered to the first dynamic PET scan and PET list mode data were rebinned into a total of 23 frames (1×15, 3×5, 3×10, 4×60, 2×150, 2×300, 4×600 and 4×300 seconds). Population based plasma-input-function in combination with a reversible two tissue compartmental model with blood volume correction (POP-IP_2T4k_VB) was used to interpolate the gap in reference tissue time activity curve (TAC).

All data was reconstructed using 3D RAMLA with a matrix size of 128×128×90 and a final voxel size of 2×2×2 mm^3^, including corrections for dead time, decay, attenuation, randoms and scatter.

T1-weighted MR scans were co-registered onto the corresponding PET images using Vinci software. Volumes of interest based on the Hammers template[26] were subsequently delineated on the MR images and superimposed on the PET scan using PVElab[27]. Binding potential (BP_ND_) and R_1_ parametric images were generated using receptor parametric mapping (RPM) with cerebellar gray matter as a reference region[28]. SUVr was generated using the 80-100 min p.i. time interval. BP_ND_, R_1_ and SUVr were extracted in the following apriori defined (three-dimensional) regions-of-interest (ROIs) in subject space: Braak I/II (entorhinal), Braak III/IV (limbic) and Braak V/VI (neocortical) regions. These ROIs align with neuropathologically defined regions[29], and are informative for tau PET in AD, as was shown previously by our group and others[30-33].

For each parameter and ROI, we calculated percentage change using the following formula (where FU and BL indicate the value of the parameter of interest (DVR (=BP_ND_+1) or SUVr) associated with the follow-up (FU) and baseline (BL) scan respectively):

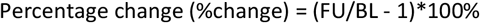

### Statistical analyses

To allow for direct comparison with SUVr values, DVR was used for all analyses.

Paired t-tests were performed to assess differences between parameters (DVR and SUVr) and time points (baseline and follow-up for DVR, SUVr and R_1_). Pearson’s correlation coefficients were computed to assess the correlation between %change SUVr and DVR (all ROIs combined). Bland-Altman analyses were performed to assess bias and agreement between %change SUVr and DVR (all ROIs combined). Analyses were performed in R software version 4.0.2. and GraphPad Prism 9.1.0.

To explore if the required sample size for (theoretical) future trials would differ when either quantitative or semi-quantitative methods are used, sample size calculations were performed using GPower v3.1.9.7. For these analyses, we used a range of 0.5% to 10% expected change in tracer retention over time, to inform on longitudinal study designs in the context of [^18^F]flortaucipir. Sample sizes were calculated for SUVr and DVR, for all three ROIs (Braak I/II, III/IV and V/VI). The differences between two dependent means (matched pairs) was calculated, with an α (error probability) of 0.05 and a power (1-β error probability) of 0.80. To adhere to the typical duration of clinical trials in AD we calculated percentage change over an 18 month period and used those standard deviations as input for the sample size calculations.

### Simulations

Simulated time activity curves were generated to assess the effect of (larger) flow changes on the accuracy of DVR/BP_ND_ and SUVr values. SRTM BP_ND_ and SUVr derived from the clinical cohort were used for this end. The outcome of the SRTM BP_ND_ results were subdivided into four conditions to assess the effects of flow changes in different binding levels. The four conditions were: SCD condition (almost no binding) and high, medium or low binding conditions for AD patients. Table-1 presents the associated parametric values for each condition used for the simulations. These values were used as reference for evaluation of the simulations and are referred as the “true” value. For each level of binding, R_1_ values were varied to mimic 5 to 25% increase or decrease in flow (in steps of 5). Efflux rate of the tracer to the blood compartment (k_2_) values were also changed accordingly. In total, there were 11 different flow conditions for each level of binding (including the reference flow condition). An average TAC of the cerebellum gray was obtained from the clinical data and was used as the reference region. For each of the flow and binding conditions, 50 target tissue TACs were generated at noise level with a coefficient of variation (COV) of 5%.

**Table-1.** “True” values (for each parameter) based on clinical data determined for each condition for the simulations. All micro/macro-parameters were estimated using simplified reference tissue model.

| | $BP_{ND}$ | $k_2$ | $R_1$ |
| --- | --- | --- | --- |
| SCD: almost no binding | 0.05 | 0.05 | 0.91 |
| AD: low binding | 0.11 | 0.04 | 0.89 |
| AD: medium binding | 0.21 | 0.03 | 0.85 |
| AD: high binding | 0.32 | 0.08 | 0.88 |

Subsequently, non-linear regression (NLR) based simplified reference tissue model (SRTM) was fitted to each of these TACs to estimate the micro/macro-parameters such as R_1_ and BP_ND_. In addition, SUVr_80-100min_ was obtained for each flow condition and binding conditions. Furthermore, the average of the 50 TACs per condition were used to assess the effect of change in R_1_ on DVR and SUVr. Simulations were performed with noise (5%) and with almost no noise (0.05%) to the TACs, using the noise model described by Yaqub et al.[34] for both the simulations. Note that 5% COV corresponded visually to noise observed in regional TACs obtained from clinical data.

SUVr values at time intervals other than 80-100 min were also obtained for each simulated flow and binding condition to evaluate the effect of the choice of uptake interval. Percentage bias for estimated parametric values (DVR and SUVr) was calculated with respect to true DVR values illustrated in Table-1. Furthermore, percentage change for each SUVr interval was calculated with respect to true DVR values and the effect of flow was also evaluated on these measures.

## RESULTS

Patient characteristics are shown in Table-2. In both AD and SCD, [^18^F]flortaucipir SUVr were higher compared to DVR for all regions and at both time points (baseline and follow-up, all p<0.001). Respective DVR, SUVr and R_1_ values are shown in Table-3 and Table-4. The percentage overestimation of SUVr relative to DVR, for all regions and at both time points, are presented in sTable-1. Annualized percentage change for DVR and SUVr are presented in sTable-2 and sTable-3.

**Table-2.**
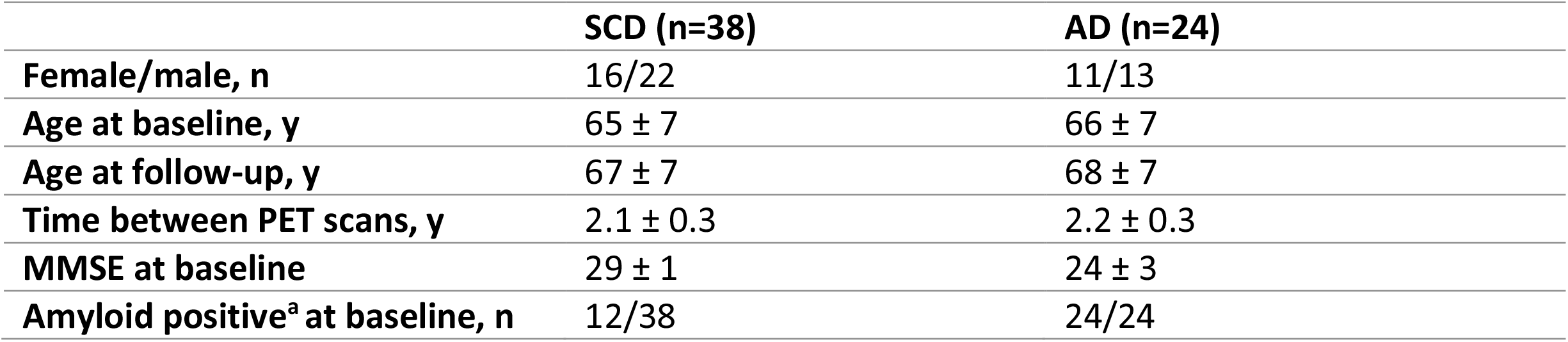
Demographics of the study population. Mean ± SD are provided, unless otherwise indicated. SCD = subjective cognitive decline, AD = Alzheimer’s disease, MMSE = mini mental state examination. ^A^SCD subjects were classified as amyloid positive as evidenced by substantial Aß pathology after SUVr_50-70min_ [^18^F]florbetapir Aß-PET scan visual assessment [20] and MCI/AD patients were classified as amyloid positive as evidenced by CSF biomarkers for AD (i.e. CSF Aß_1-42_ < 813 ng/L [21]) and/or a positive Aß-PET ([^18^F]PiB or [^18^F]florbetaben) scan by visual assessment [22, 23].

**Table-3.** [^18^F]flortaucipir DVR, SUVr and R_1_ values for SCD subjects. Mean ± SD are provided. DVR = BP_ND_+1.

| SCD (n=38) | DVR |  |  | SUVR<sub>80-100</sub> |  |  | R<sub>1</sub> |  |  |
| --- | --- | --- | --- | --- | --- | --- | --- | --- | --- |
|  | BL | FU | %change | BL | FU | %change | BL | FU | %change |
| Braak I/II | 1.039 (0.121) | 1.066<sup>z</sup> (0.133) | 2.56* (2.85) | 1.134<sup>a</sup> (0.159) | 1.154<sup>ay</sup> (0.158) | 1.85 (3.27) | 0.708 (0.041) | 0.714 (0.049) | 0.74 (3.96) |
| Braak III/IV | 1.045 (0.075) | 1.075<sup>z</sup> (0.098) | 2.82 (2.54) | 1.102<sup>a</sup> (0.103) | 1.130<sup>az</sup> (0.118) | 2.47 (2.64) | 0.836 (0.036) | 0.842 (0.043) | 0.79 (2.75) |
| Braak V/VI | 1.042 (0.057) | 1.067<sup>z</sup> (0.076) | 2.33 (2.77) | 1.076<sup>a</sup> (0.077) | 1.096<sup>az</sup> (0.093) | 2.17 (3.29) | 0.926 (0.043) | 0.930 (0.049) | 0.47 (2.67) |

DVR vs. SUVR: <sup>a</sup>p<0.001 | BL vs. FU: <sup>x</sup>p<0.05, <sup>y</sup>p<0.01, <sup>z</sup>p<0.001 | %change DVR vs. %change SUVR: \*p<0.05

### Differences in [^18^F]flortaucipir DVR, SUVr and R_1_

#### SCD subjects

DVR increased at follow-up in all regions (all p<0.001), with largest increases found in the Braak III/IV (from 1.045 to 1.075, 2.82±2.54%) and Braak I/II region (from 1.039 to 1.066, 2.56±2.85%), and smallest increases in the Braak V/VI region (from 1.042 to 1.067, 2.33±2.77%) (Table-3 and Figure-1a&2a). SUVr also significantly increased at follow-up in all regions (all p<0.003). Largest increases were found in the Braak III/IV (from 1.102 to 1.130, 2.47±2.64%) and Braak V/VI region (from 1.076 to 1.096, 2.17±3.29%), and smallest increases in the Braak I/II region (from 1.134 to 1.154, 1.85±3.27%) (Table-3 and Figure-1c&2c). Percentage change was significantly lower for SUVr compared to DVR in the Braak I/II region (SUVr 1.85±3.27% vs DVR 2.56±2.85%, p=0.048). Braak III/IV and V/VI regions did not show any statistically significant differences between percentage change in DVR and SUVr (Table-3 and Figure-3). Taking all regions together, the correlation coefficient between %change in SUVr and DVR was 0.83 (p<0.001), and the bias as provided by Bland-Altman analysis was 0.41±1.72 (Figure-4a&c). For R_1_, no significant decreases at follow-up were found in any region (Table-3).

**Figure-1.**
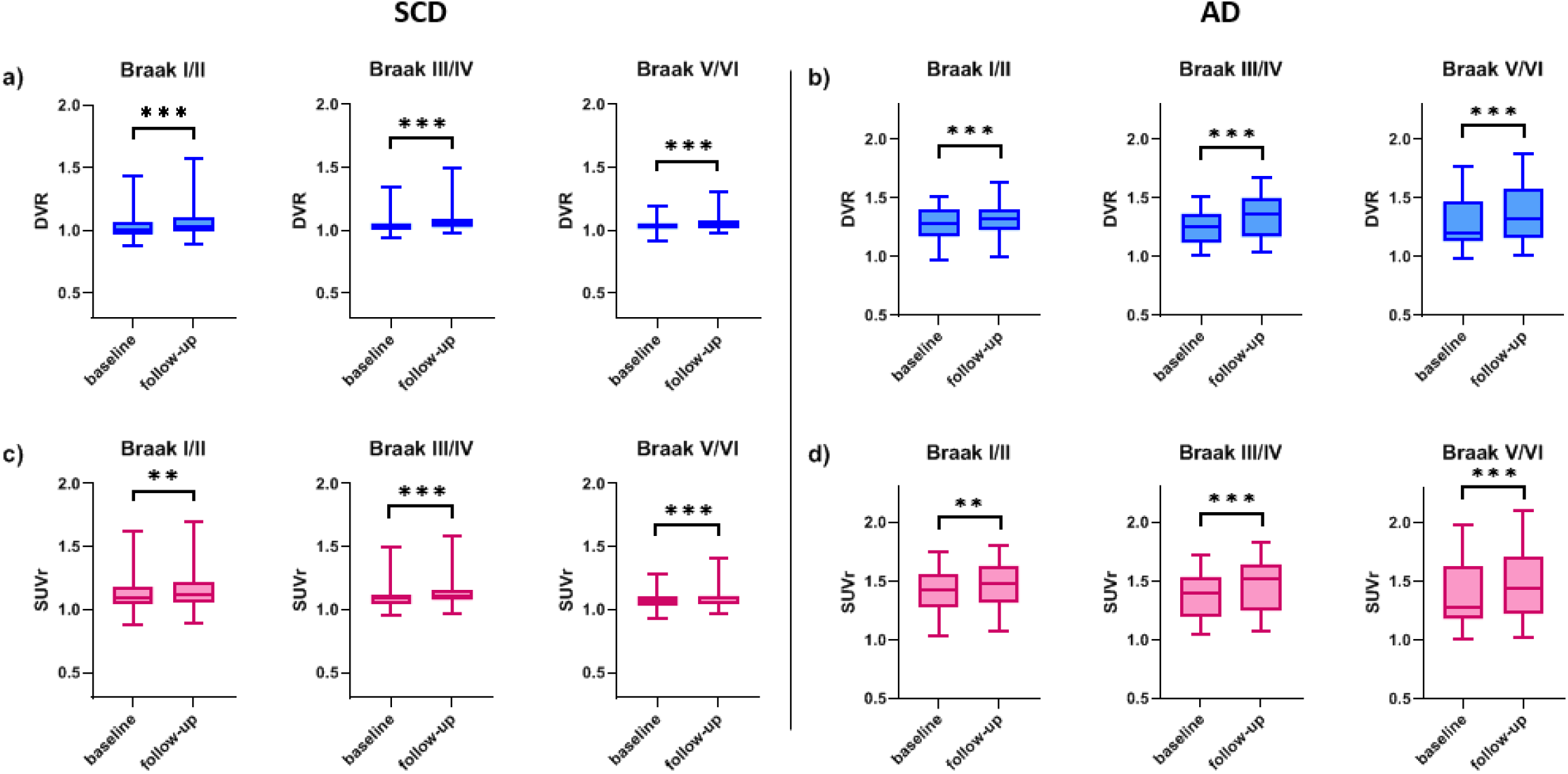
Boxplots of regional DVR in **(a)** SCD and **(b)** AD. Boxplots of regional SUVr_80-100_ in **(c)** SCD and **(d)** AD. **p<0.01, ***p<0.001

**Figure-2.**
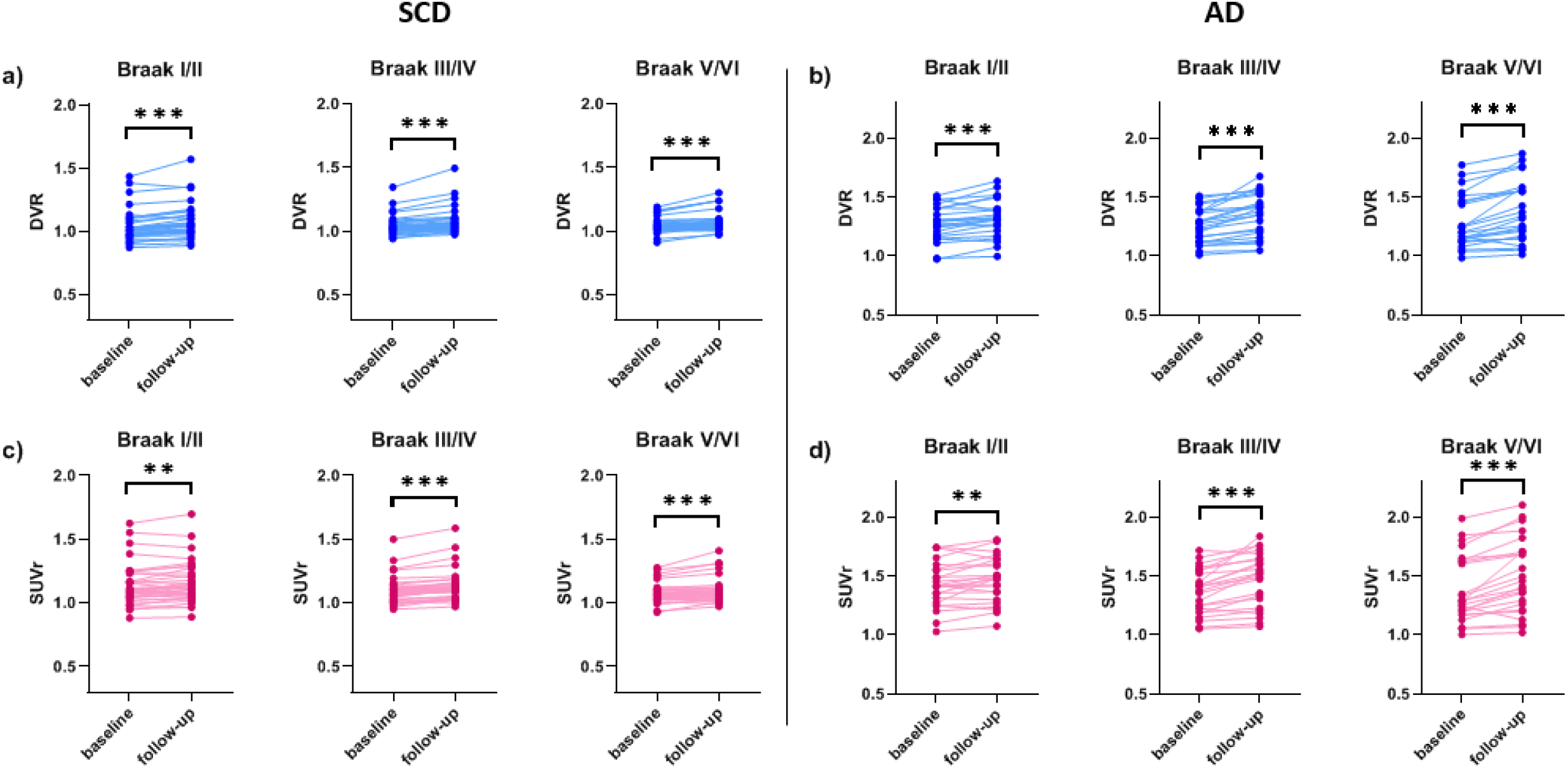
Spaghetti plots of regional DVR in **(a)** SCD and **(b)** AD. Spaghetti plots of regional SUVr_80-100_ in **(c)** SCD and **(d)** AD. **p<0.01, ***p<0.001

**Figure-3.**
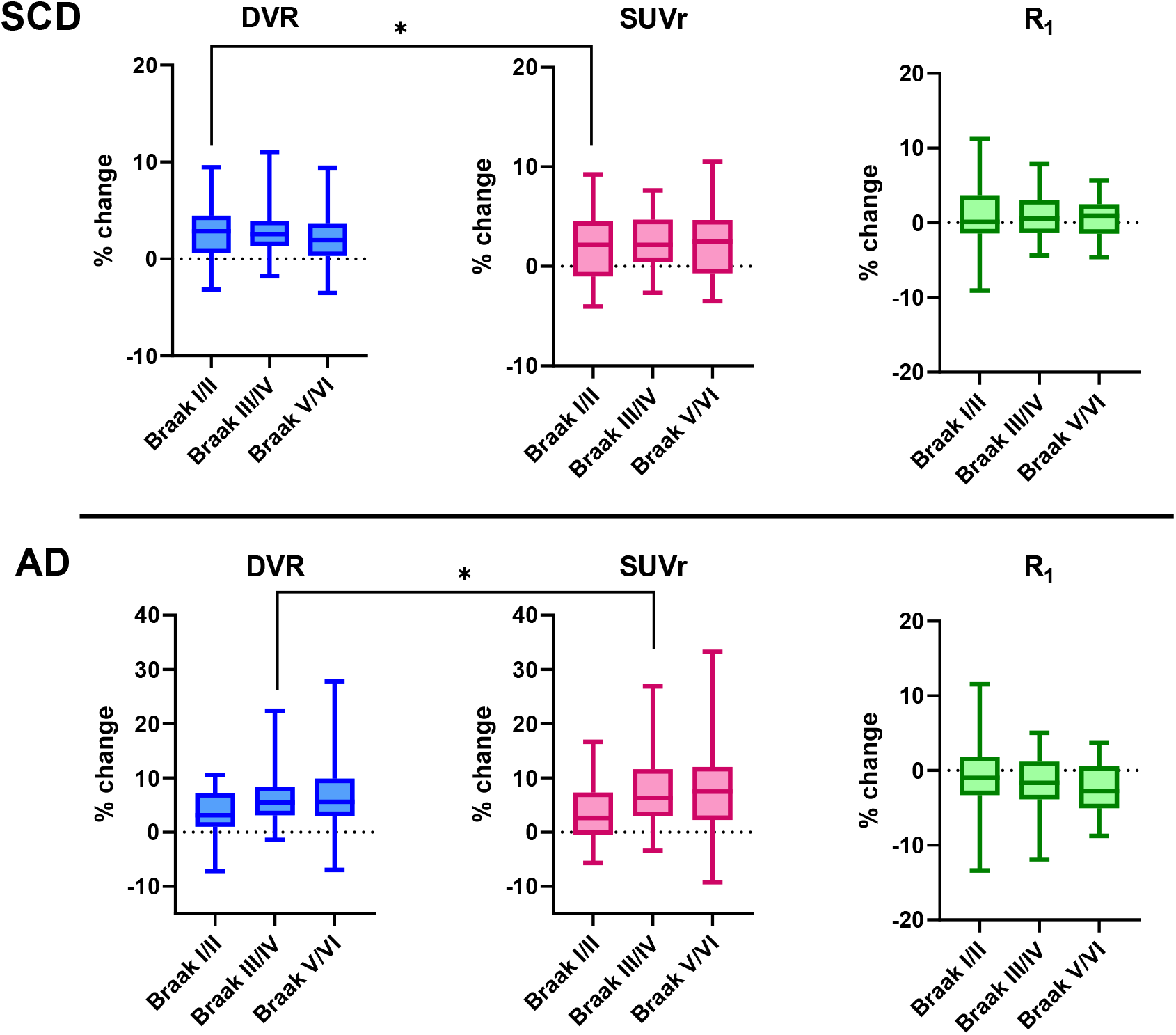
Regional percentage changes in DVR, SUVr_80-100_ and R_1_ for SCD subjects and AD patients. *p<0.5, **p<0.01, ***p<0.001

**Figure-4.**
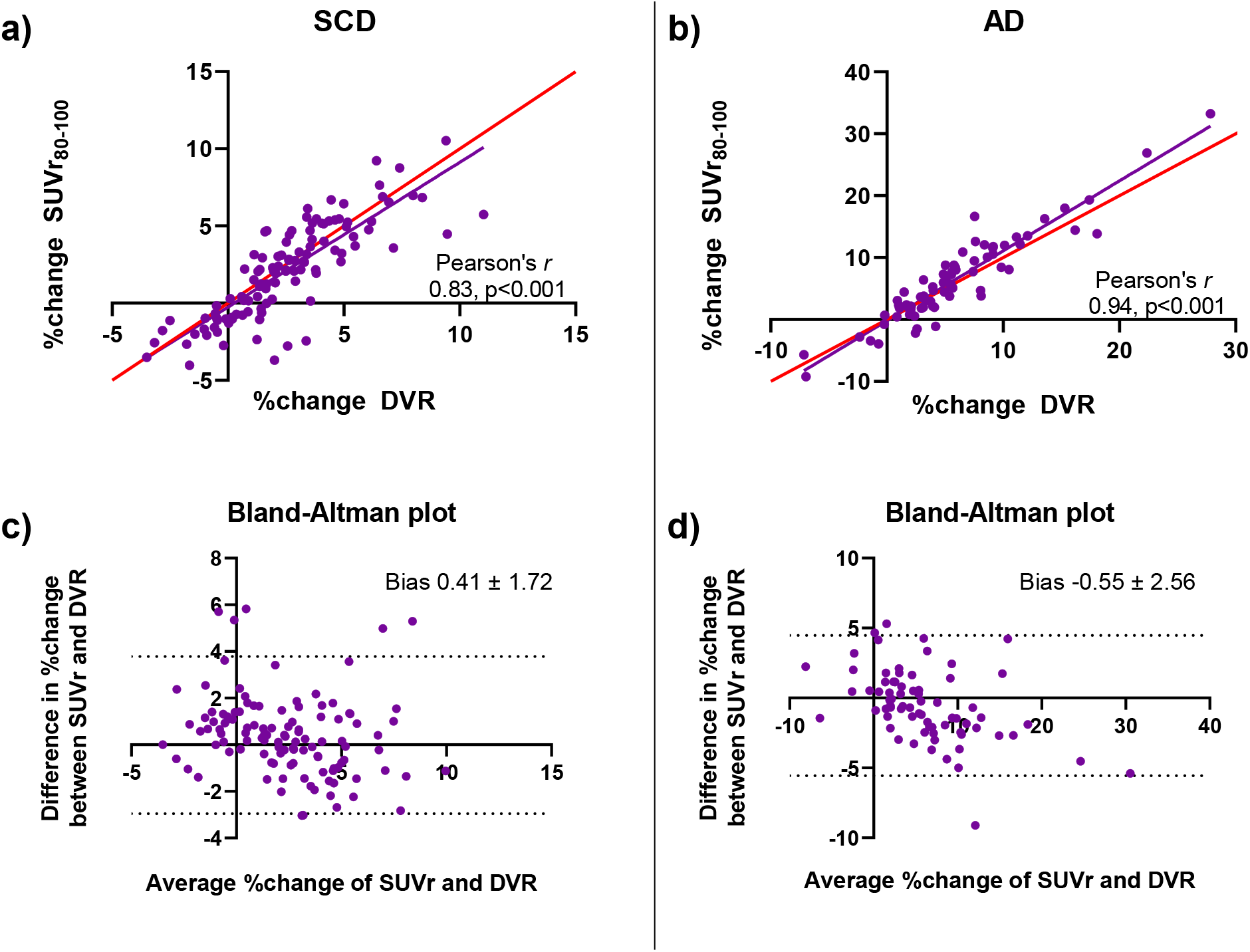
Correlation plot of %change in DVR vs SUVr_80-100_ in **(a)** SCD and **(b)** AD, in which the red line represents the line of identity. Bland-Altman plot of %change in DVR vs SUVr_80-100_ in **(c)** SCD and **(d)** AD.

#### AD patients

DVR increased at follow-up in all regions (all p<0.001), with largest increases found in the Braak V/VI (from 1.284 to 1.379, 7.25±6.85%) and Braak III/IV region (from 1.256 to 1.341, 6.61±5.63%), and smallest increases in the Braak I/II region (from 1.277 to 1.321, 3.48±4.16%) (Table-4 and Figure-1b&2b). SUVr also increased at follow-up in all regions (all p<0.009). Like for DVR, largest increases were found in the Braak V/VI (from 1.382 to 1.495, 8.21±8.03%) and Braak III/IV region (from 1.367 to 1.471, 7.52±6.66%), and smallest increases in the Braak I/II region (from 1.426 to 1.470, 3.25±5.26%) (Table-4 and Figure-1d&2d). Percentage change was higher for SUVr compared to DVR in the Braak III/IV region (SUVr 7.52±6.66% vs DVR 6.61±5.63%, p=0.047). No statistically significant differences between percentage change of SUVr and DVR were found for any other region (Table-4 and Figure-3). Taking all regions together, the correlation coefficient between %change in SUVr and DVR was 0.94 (p<0.001) and the bias as provided by Bland-Altman analysis was -0.55±2.56 (Figure-4b&d). For R_1_, significant decreases at follow-up were found in Braak III/IV (from 0.835 to 0.821, - 1.62±3.71%, p=0.040) and V/VI regions (from 0.904 to 0.883, -2.28±3.67%, p=0.003), but not in the Braak I/II region (from 0.713 to 0.706, -0.87±5.26%, p=0.372) (Table-4).

**Table-4.**
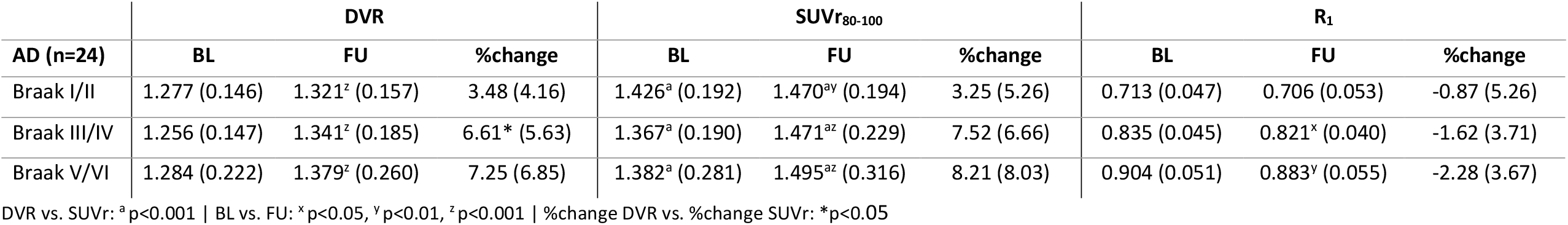
[^18^F]flortaucipir DVR, SUVr and R_1_ values for AD patients. Mean ± SD are provided. DVR = BP_ND_+1.

### Sample size calculations

Large differences in required sample sizes were observed for small effect sizes, with largest differences between methods in the AD group (Table-5). However, with larger effect sizes (in line with expectations in clinical trials) differences in sample size between both methods became negligible for both SCD and AD (Table-5).

**Table-5.** Required sample sizes (n) for [^18^F]flortaucipir for different effect sizes.

| SCD |  |  |  |  |  |  |  |  |  |  |  |  |
| --- | --- | --- | --- | --- | --- | --- | --- | --- | --- | --- | --- | --- |
| Expected change |  | 0.5% | 1% | 2% | 3% | 4% | 5% | 6% | 7% | 8% | 9% | 10% |
| DVR | Braak I/II | 141 | 37 | 11 | 7 | 5 | 4 | 4 | 3 | 3 | 3 | 3 |
|  | Braak III/IV | 117 | 31 | 10 | 6 | 5 | 4 | 4 | 3 | 3 | 3 | 3 |
|  | Braak V/VI | 131 | 34 | 11 | 6 | 5 | 4 | 4 | 3 | 3 | 3 | 3 |
| SUV<sub>r80-100</sub> | Braak I/II | 180 | 47 | 14 | 8 | 6 | 5 | 4 | 4 | 3 | 3 | 3 |
|  | Braak III/IV | 119 | 32 | 10 | 6 | 5 | 4 | 4 | 3 | 3 | 3 | 3 |
|  | Braak V/VI | 177 | 46 | 13 | 8 | 5 | 5 | 4 | 4 | 3 | 3 | 3 |
| AD |  |  |  |  |  |  |  |  |  |  |  |  |
| Expected change |  | 0.5% | 1% | 2% | 3% | 4% | 5% | 6% | 7% | 8% | 9% | 10% |
| DVR | Braak I/II | 223 | 58 | 16 | 9 | 6 | 5 | 4 | 4 | 4 | 3 | 3 |
|  | Braak III/IV | 324 | 83 | 23 | 12 | 8 | 6 | 5 | 4 | 4 | 4 | 4 |
|  | Braak V/VI | 497 | 126 | 33 | 16 | 10 | 8 | 6 | 5 | 5 | 4 | 4 |
| SUV<sub>r80-100</sub> | Braak I/II | 391 | 100 | 27 | 13 | 9 | 7 | 5 | 5 | 4 | 4 | 4 |
|  | Braak III/IV | 446 | 113 | 30 | 15 | 10 | 7 | 6 | 5 | 5 | 4 | 4 |
|  | Braak V/VI | 641 | 162 | 42 | 20 | 13 | 9 | 7 | 6 | 5 | 5 | 4 |

### Simulations

Simulations with 5% COV showed similar results as the simulated TACs obtained with almost no noise (0.05% COV). Therefore, only the results from TACs with 5% COV were reported in the results to mimic real cohort data.

Simulations revealed that in the SCD (almost no binding) condition and low binding AD patient condition, an inverse relation was observed i.e. with increasing flow a decreasing bias for SUVr (with respect to true DVR) was observed (Figure-5). A similar behaviour was also observed in case of the medium binding AD patient condition, however to a lesser extent. Although, in the high binding condition for AD patients, relatively a smaller effect of flow was observed on the SUVr, implying that SUVr values remained relatively constant irrespective of the change in flow. In case of DVR, no effect of flow was observed with any of the conditions (Figure-5).

**Figure-5.**
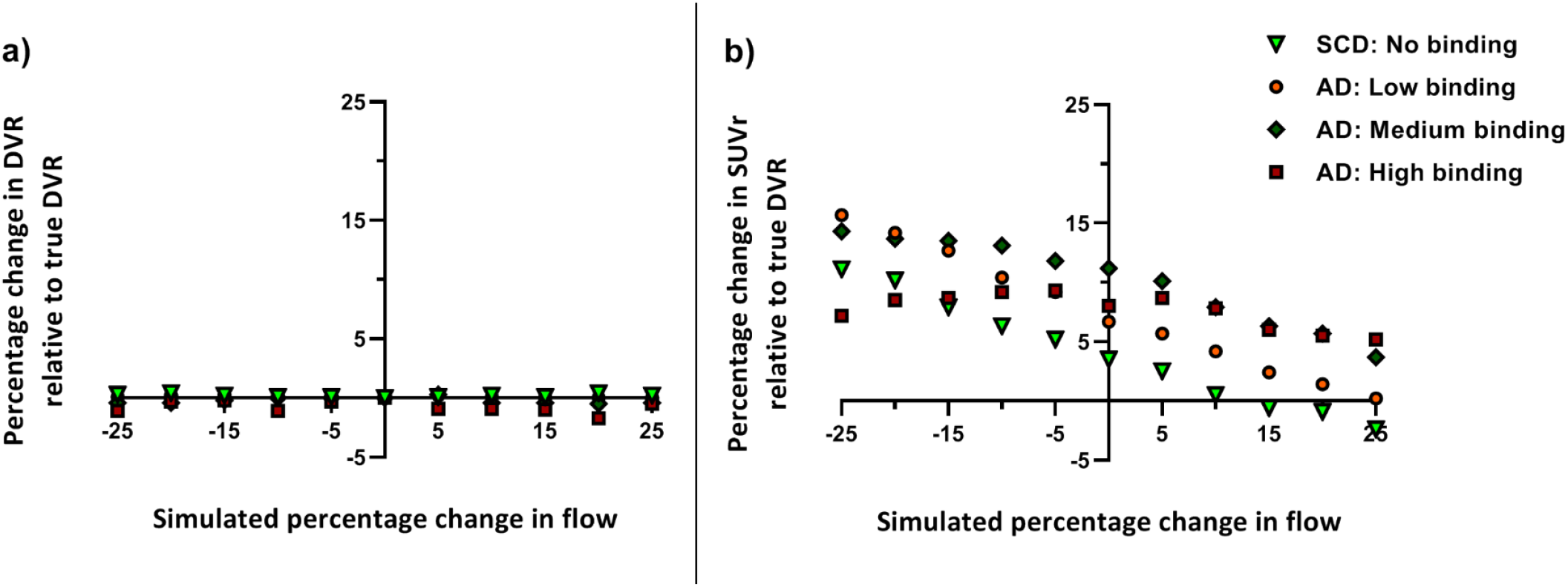
Percentage change in **(a)** SUVr_80-100_ and **(b)** DVR relative to the true DVR values as a function of simulated flow changes for each binding condition.

Based on simulations, percentage bias in SUVr with respect to the true DVR varied with the choice of SUVr uptake interval and the underlying binding condition (Figure-6). In general, SUVr overestimated DVR for all simulated R_1_ conditions from 80 min p.i., however, the impact of the change in flow on the directionality of the bias seems also to vary with respect to the choice of SUVr uptake time interval (Figure-6).

**Figure-6.**
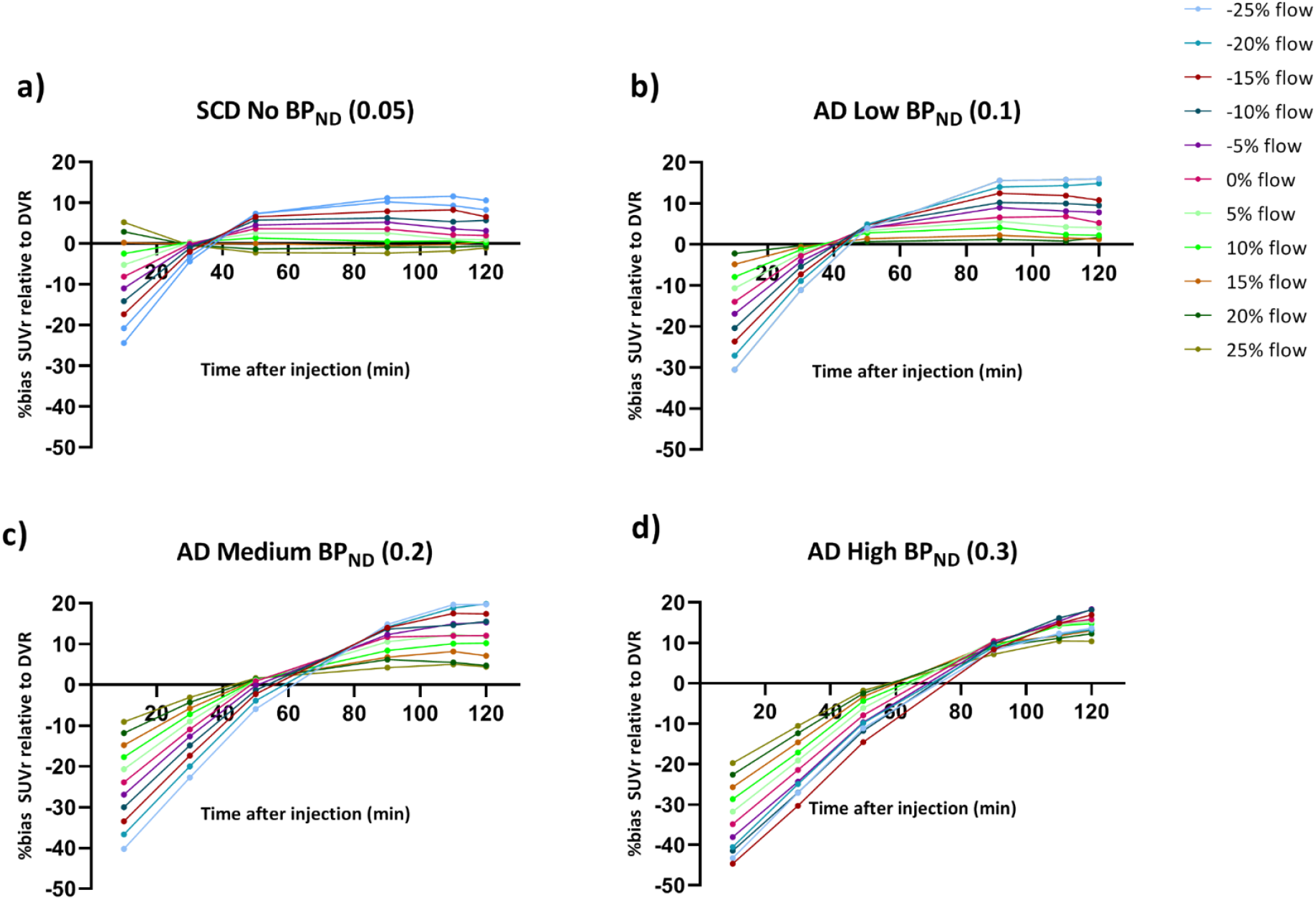
Percentage bias in SUVr relative to the true DVR values as a function of SUVr time intervals for simulated flow condition for **(a)** SCD (almost no binding condition) **(b)** AD with low binding condition **(c)** AD with medium binding condition and **(d)** AD with high binding condition. Legend in the figure illustrates different flow conditions (increase or decrease in flow 0 – 25%).

## DISCUSSION

In this study, we compared assessment of changes in [^18^F]flortaucipir specific binding using a static (SUVr) and dynamic (DVR (= BP_ND_+1)) analytic method. In a two-year longitudinal study, [^18^F]flortaucipir did not show large differences in change over time between DVR and SUVr. This suggests that SUVr provides an accurate estimate of specific binding in this setting (where changes in blood flow were small). However, simulations demonstrated marked differences between DVR and SUVr when large(r) changes in blood flow (R_1_) were introduced. In addition, these differences between DVR and SUVr were shown to be dependent on the underlying level of tau pathology as they were most prominent in individuals with a high tau load. This is especially important for trials using pharmacotherapeutic interventions, since pharmacotherapeutics could potentially induce large flow changes.

The lack of significant differences between SUVr and DVR in the longitudinal setting (observational dataset), could be explained by the relatively small or nonexistent flow/R_1_ differences in this cohort. Therefore, simulations were performed to investigate the degree of impact on the accuracy of both methods/parameters (SUVr and SRTM DVR) with introducing large(r) changes in flow/R_1_. As can be seen from Figure-5, not only the bias with SUVr relative to the true DVR was different for each flow conditions but also was influenced with underlying tau load. Depending on the underlying tau load/binding, regional changes in flow, resulted in variable changes in SUVr (decreasing bias in case of low tau load/binding or constant bias for high tau load/binding), which was not the scenario with DVR. Similar findings were earlier observed using [^18^F]Cyclofoxy[35]. Carson et al. concluded that SUVr can overestimate specific binding substantially due to the sensitivity to differences in clearance rate. They also observed that the bias was different for high- and low-binding areas, which is in line with our results from the simulations. Another study also observed that kinetic methods such as RPM2 and reference Logan showed relatively stable estimates of [^11^C]-PiB binding in AD patients over time, while SUVr showed larger variability between subjects[6]. The authors concluded that this variability could be related to changes in R_1_ over time, since it was observed in the study cohort that R_1_ decreased in AD patients over time.

As can be observed in Figure-6, the choice of SUVr time interval also effected the accuracy, which was also different for different binding conditions. A previous study found large positive biases for SUVr using different time intervals when compared to dynamic methods[28]. Furthermore, Golla et. al. observed that the bias in SUVr for a specific uptake interval is not constant, but is dependent on the underlying tau load and the choice of SUVr uptake interval. This has important implications, since as uptake time intervals for static protocols are often not strictly enforced, leading to deviations in uptake time between various static, longitudinal scans being very common. These discrepancies will increase variability and uncertainty, which will lead to increases in required sample sizes for SUVr. Differing underlying tau load in the sample studied will only increase the bias in SUVr further. It is worth noting that, in the current study, SUVr was extracted from the dynamically acquired data. In addition, uptake time was strictly enforced in the context of the two scanning sessions within the dynamic protocol. For both these reasons, SUVr in this study was not affected by deviations in uptake time.

The discrepancies between methods using simulations have important implications on longitudinal [^18^F]flortaucipir studies and intervention studies. Our findings imply that SUVr should not be used when variations in blood flow are expected. Particularly if the study dataset comprises of subjects with varying tau load, since from the simulation it was clear that the effect of flow on SUVr with such a dataset might not be unidirectional. Therefore, dynamic scanning protocols and fully quantitative data analysis methods are essential for accurate quantification of longitudinal tau PET studies, especially for studies with disease-modifying treatments aiming to lower the amount of tau in the brain directly or indirectly, as it had been demonstrated that pharmacotherapeutic interventions in AD could induce changes in cerebral blood flow[36, 37]. However, the scan duration for [^18^F]flortaucipir more than doubles when using fully quantitative dynamic acquisition, even with a shortened dynamic protocol with a total scanning duration of 50 min instead of 110 min[25]. Therefore, a consideration to address when using repeated 100/130 min dynamic scans is potential selection bias, because severely affected patients might not be able to undergo such a demanding procedure. In case of moderate to severe AD patients this is indeed debatable. Although, pharmacotherapeutic trials currently show a shift in target-population, primarily including patients with mild, prodromal or preclinical autosomal-dominant AD. Those patients are most likely to tolerate the dynamic scan procedures.

The present study demonstrated no major differences in the percentage change between [^18^F]flortaucipir DVR and SUVr values in a two-year observational study. Congruently, sample size calculations based on these data to inform future trials showed negligible differences between methods. These findings are not completely in line with previous amyloid PET studies showing that dynamic scanning outperformed static protocols. There are several reasons that might explain this. First, it has previously been reported that accumulation of tau pathology is a slowly developing process, with annual percentage changes of about 0.5-3% in amyloid positive cognitively unimpaired subjects and up to 3-10% in amyloid positive cognitively impaired subjects (based on SUVr)[38-41]. The annual percentages change in the present study are in general comparable in the SCD subjects (on average 1.08% SUVr and 1.28% DVR), and slightly lower in AD subject (2.73% SUVr and 2.52% DVR). The test-retest repeatability of [^18^F]flortaucipir, as reported previously[42], lies around 1.98% (0.78–3.58) for DVR and 3.05% (1.28–5.52) for SUVr_(80-100min)_. Although the test-retest repeatability was significantly better for DVR[42], annual percentage changes as found in the present study still fall within one standard deviation of the test-retest repeatability for both DVR and SUVr, suggesting that observed changes might be too small to detect differences between analytic methods. Second, an important cause of the longitudinal difference between DVR and SUVr is the differing blood flow dependence[6, 35]. Although for this study, R_1_ values decreased at follow-up in AD, decreases were relatively small (−1.59% on average), and most likely too small to lead to statistically detectable differences between methods. For comparison, a previous study showed marginal statistical differences between [^11^C]PiB DVR and SUVr when R_1_ decreased on average 4.6% (from 0.87 to 0.83) in AD[6], which is a larger change compared to the present study. Third, the study population might have influenced the results. Previous findings showed most pronounced differences between analytic methods in AD patients rather than MCI or control subjects[6]. In the present study, subjects with MCI due to AD and probable AD dementia were grouped into one AD group. Fourth, differences with respect to tracer target (amyloid plaques vs paired helical filament tau), isotope (^11^C vs ^18^F) and pharmacokinetic behavior might have introduced differences inherent to differences in results.

In conclusion, dynamic scanning protocols and fully quantitative data analysis methods must be preferred for accurate quantification of longitudinal [^18^F]flortaucipir imaging studies where large(r) flow changes in the brain (such as in later disease stages and/or pharmacotherapeutic interventions) are expected. Usage of semi-quantitative methods in such conditions, such as SUVr, carries the inherent risk that potential effective therapeutic interventions are discarded, especially when expected effect sizes are small.

## Data Availability

Anonymized data used in the present study may be available upon request to the corresponding author.

## Compliance with Ethical Standards

### Funding

This study was funded by a ZonMW Memorabel grant.

### Ethical approval

All procedures performed in studies involving human participants were in accordance with the ethical standards of the institutional and/or national research committee and with the 1964 Helsinki declaration and its later amendments or comparable ethical standards.

### Informed consent

Informed consent was obtained from all individual participants included in the study.

### Disclaimer

Denise Visser and Hayel Tuncel contributed equally to this work.

## Acknowledgments

We kindly thank all participants for their contribution. We thank Ronald Boellaard for sharing his knowledge and thoughts about the project. Research of Amsterdam Alzheimer Center is part of the Neurodegeneration program of Amsterdam Neuroscience. The Amsterdam Alzheimer Center is supported by Alzheimer Nederland and Stichting VUmc funds. [^18^F]Flortaucipir PET scans were made possible by Avid Radiopharmaceuticals Inc.

