## Supplementary Material for "Longitudinal tau PET using [^18^F]flortaucipir: Comparison of (semi)quantitative parameters"

† both authors contributed equally

Corresponding author:

**sTable-1. Percentage overestimation of $SUVr_{80-100}$ relative to DVR.** Mean ± SD are provided.

| | SCD | | AD | |
|---|---|---|---|---|
| | **BL** | **FU** | **BL** | **FU** |
| **Braak I/II** | 8.95 (3.92) | 8.16 (3.39) | 11.49 (3.46) | 11.18 (3.21) |
| **Braak III/IV** | 5.34 (3.02) | 4.97 (2.67) | 8.59 (3.04) | 9.47 (3.29) |
| **Braak V/VI** | 3.19 (2.35) | 3.00 (1.98) | 7.08 (3.59) | 7.97 (3.47) |

**sTable-2. Annualized %change for [$^{18}$F]flortaucipir DVR and $SUVr_{80-100}$ in SCD subjects.** Mean ± SD are provided. DVR = $BP_{ND}$+1.

| SCD (n=38) | Annualized %change DVR | Annualized %change $SUVr_{80-100}$ |
|---|---|---|
| **Braak I/II** | 1.27 (1.44) | 0.92 (1.62) |
| **Braak III/IV** | 1.41 (1.31) | 1.23 (1.32) |
| **Braak V/VI** | 1.16 (1.38) | 1.08 (1.61) |

**sTable-3. Annualized %change for [$^{18}$F]flortaucipir DVR and $SUVr_{80-100}$ values in AD patients.** Mean ± SD are provided. DVR = $BP_{ND}$+1.

| AD (n=24) | Annualized %change DVR | Annualized %change $SUVr_{80-100}$ |
|---|---|---|
| **Braak I/II** | 1.54 (1.77) | 1.42 (2.35) |
| **Braak III/IV** | 2.86 (2.13) | 3.23 (2.51) |
| **Braak V/VI** | 3.15 (2.65) | 3.53 (3.01) |
